# *“It is bad because it limits capacity building here back at home”* Genetic and genomic researchers’ perspectives on biological sample sharing in collaborative research

**DOI:** 10.1101/2022.10.19.22281283

**Authors:** David Kaawa-Mafigiri, Deborah Ekusai- Sebatta, Ian Munabi, Erisa Sabakaki Mwaka

## Abstract

Numerous ethical, legal and social issues arise with biological sample sharing. The study explored the perspectives of genetic/genomic researchers on the sharing of biological samples in international collaborative research. Qualitative in-depth interviews were conducted with 15 researchers. Participants expressed positive attitudes towards biobanking and appreciated the benefits of cross-border sharing of biological samples but noted that this practice had adversely affected local capacity building efforts. There was limited understanding of the ethico-regulatory frameworks governing sample sharing. Researchers emphasized the importance of respecting cultural values in biobanking research. Issues concerning poor governance and inequitable benefit sharing were also raised. There is a need for fair and equitable international collaborations where all researchers are treated with respect and as equal partners.

## Introduction

The need for scientific advancement has made the use of human biological samples and research data for biomedical research an area of high interest for various stakeholders (Hansson, 2009). Over the years, large quantities of biological samples have been shipped from low- and middle-income countries (LMIC) to developed countries for various reasons including the lack of capacity for storage, lack of appropriate laboratory infrastructure, and lack of technical capacity (Matandika et al., 2020; Staunton & Moodley, 2013). This has also been attributed to weak regulatory systems (de Vries et al., 2017; Whitney et al., 2008). Empirical evidence has showed that the ethico-regulatory frameworks regulating the acquisition, storage, use and transfer of biological materials in several African countries are inadequate, differ substantially and may conflict across borders (de Vries et al., 2017; Staunton & Moodley, 2013).

With increasing globalization, there is an increase in the importance of ethical, legal and social implications of research involving the use of biological samples and associated data on participant autonomy, informed consent, confidentiality, privacy, reuse and ownership, return of results, data sharing, and benefit sharing with communities (Barnes & Heffernan, 2004; Budimir et al., 2011; Cambon-Thomsen, Rial-Sebbag, & Knoppers, 2007; Hansson, 2011).

Globally, several studies have reported a positive attitude of various stakeholders towards genomic research and biobanking research (Lemke, Halverson, & Ross, 2012; Lemke, Wolf, Hebert-Beirne, & Smith, 2010; McGuire, Hamilton, Lunstroth, McCullough, & Goldman, 2008; Pentz, Billot, & Wendler, 2006; Trinidad et al., 2010; Vermeulen, Schmidt, Aaronson, Kuenen, & van Leeuwen, 2009). Studies amongst research participants have reported varying attitudes towards storage and sharing of biological samples for future research. Whereas some believed that it is unethical to conduct future studies on stored biological samples (Al-Ebbini, Khabour, Alzoubi, & Alkaraki, 2021), others had no concerns (Verstuyft et al., 2018).

In sub-Saharan Africa, studies have reported mixed attitudes towards biobanking research. Whereas some studies have reported support for sharing of biological samples in collaborative research (Matandika et al., 2020; Mweemba et al., 2020; Tindana, Molyneux, Bull, & Parker, 2014), others have reported reluctance to share (Moodley & Singh, 2016; Singh & Moodley, 2021). Further, there is mistrust between African researchers and their international collaborators because of the historical exploitative research that has been conducted in low resource settings (Matandika et al., 2020; Moodley & Singh, 2016; Munung, Mayosi, & De Vries, 2018; Sathar, Dhai, & van der Linde, 2014). Concerns have also been raised about the lack of appropriate mechanisms to safeguard the interests of sample donors and their communities, and local investigators (Cambon-Thomsen et al., 2007; de Vries et al., 2015).

Research in Uganda is guided by the National Guidelines for Research involving Humans as Research Participants (Uganda National Council for Science and Technology, 2014) and the National Research Biobanking Guidelines (Uganda National Council for Science and Technology, 2021), that provide guidance on the acquisition, storage, and the transfer of biological samples. The transfer of biological samples across Ugandan borders can only be done with clearance from Uganda National Council for Science and Technology (UNCST) and a material transfer agreement (MTA) must be in place. According to these guidelines, biological samples can only be exported after demonstrating that in-country capacity to perform the required analyses does not exist or is inadequate. Biological samples can also be exported for quality control and reference purposes. It is important to note that the Uganda national ethics guidelines lack some key procedural details and may need improvement (Mahomed, 2020; Nnamuchi, 2016).

Uganda is a recipient of several Human Heredity and Health in Africa (H3Africa) projects that have contributed to exponential capacity building for genomics and biobanking research (H3Africa, 2014). H3Africa is a consortium of researchers from Africa that was established in 2012 with a goal of empowering Africa researchers in genomic sciences and biobanking, establishing and nurturing effective collaborative partnerships among African researchers based on the continent and generating valuable data that could be used to improve global health (H3Africa). There has been exponential increase in genomic and biobanking research on the African continent particularly under the auspices of the H3Africa initiative. The H3Africa Consortium has a biorepository program that is managing the collection of DNA and other biological materials from 22 African countries, and these are stored in regional biorepositories in Uganda, Nigeria and South Africa (H3Africa). H3Africa is currently processing samples and data for more than 70,000 participants across the African Continent (H3Africa). Specific to Uganda, is the Integrated Biorepository of H3Africa Uganda (IBRH_3_AU) that is custodian to 300,000 biological samples from more than 100,000 participants and has contributed immensely to regional biobank governance (“Integrated Biorepository of H3Africa Uganda (IBRH3AU) Biospecimen Catalogue,” 2022). We therefore anticipate continued growth of biobanking in Africa.

There are several negative precedents in sub-Saharan Africa that informed this study including 1) the alleged scandal involving the Wellcome Trust Sanger Institute that proposed to commercialize a gene chip without proper legal agreements with partner institutions and the consent of thousands of African people, whose donated DNA and data were used to develop the chip (“Major U.K. genetics lab accused of misusing African DNA,” 2019); 2) Zambia’s National Health Research Act (2013) which outrightly prohibits broad consent and the collection of biological samples for unspecified future research (Zambia, 2013), and yet a survey of societal views on this prohibition after passing the law indicated that majority of respondents preferred retaining the option of ‘broad consent’(Mweemba et al., 2019); and 3) the Malawi national ethics guidelines for genetics research that prohibit the storage and future use of biological samples for unspecified research (“Policy requirements, procedures and guidelines for the conduct and review of human genetic research in Malawi,” 2012). These developments, particularly the handling of the Wellcome Sanger situation created a lot of mistrust and suspicion among various African stakeholders. A South African University even demanded that the DNA samples that Wellcome Trust Sanger Institute wanted to commenrcialise without consent be returned to South Africa (Blakeley, 2019; Moodley & Kleinsmidt, 2020; Njilo, 2019; Singh & Moodley, 2021). This paper explores the perspectives of genetic/genomic researchers on biological sample sharing in collaborative research. As LMIC governments and scientific communities are encouraged to develop and implement policies to guide collaborative biobanking research, it is important to understand the perspectives and experiences of researchers and other stakeholders on biological sample sharing and reuse. Understanding and addressing issues within the research community that influence biological sample sharing is important during the development and implementation of biobanking policies. The findings of this study may not only contribute to the development of locally contextualized policies but may also lead to better compliance.

## Materials and Methods

As part of a bigger on-going mixed methods study exploring the perceptions and experiences of various stakeholders on the informed consent process for genetic/genomic research in Uganda (Mwaka et al., 2021) qualitative data was collected and analyzed to identify genetic/genomic researchers’ perspectives on biological sample data sharing in collaborative research.

### Study setting and participants

The study was conducted at Makerere University College of Health Sciences (MakCHS), one of the nine constituent colleges at Makerere University in Uganda. All participants were researchers actively involved in genetic/genomic research in Uganda and affiliated to Makerere University College of Health Sciences (MakCHS). Participants were principal investigators of protocols involving host genomics and genetics research that were approved by Uganda National Council for Science and Technology (UNCST) for the period 2012-2017. UNCST provides regulatory oversight of all research activities in the country; and per local regulations, all protocols approved by accredited research ethics committees are submitted to UNCST for approval and registration. We searched archived research protocols approved by UNCST for the period 2012-2017. Only investigators based at MakCHS and its affiliate research institutes were eligible. A list of 23 investigators was generated and all were invited to participate but only 15 consented and participated in the study, of which three were H3Africa principal investigators. The number of researchers conducting genetics and genomic research at MakCHS is not known. However, it is important to note that there are several masters and PhD level scientists that are in training in genetic science and bioinformatics, mainly sponsored by the H3Africa initiative (H3Africa).

### Data collection

Fifteen qualitative in-depth interviews were conducted between February to June 2019 focusing on knowledge, perceptions and experiences of genetics and genomics researchers on the storage and future use of biological materials for research. Twelve of the researchers were male. All participants were purposively selected as they were conducting genetic/genomic research at MakCHS. The interviews focused on 4 main domains for analysis: 1) opinion on the collection of BM for reuse; 2) opinion on the BM export/transfer and regulation of biobanking research; 3) challenges faced by local researchers in collaborative biobanking research; and 4) possible solutions to improve/realize outcomes of biological sample and associated data sharing.

Interviews, lasting between 45-60 minutes, were conducted by a team of four researchers comprising of a qualified, trained social scientist (DES), a research assistant who was also a graduate student of bioethics, one medical anthropologist (DKM) with expertise in qualitative research methods, and one Bioethicist (ESM) with training in medicine. The same team of four conducted all interviews to ensure consistency. Prior to the start of the study, the research team was trained on the protocol to ensure that they internalized it well. Data were collected using an in-depth interview guide that was developed by ESM, DKM and DES, and explored perspectives of genetic/genomic researchers on biological sample sharing in collaborative research. The interview guide was piloted and revised prior to the full data collection process. All interviews were conducted in English, and audio recorded using an Olympus model digital recorder. Audio data was later transcribed verbatim using MS Word processor. Audio data was supplemented by notes taken during the interviews. Debrief meetings were held by the research team at the end of each interview to check for completeness and review preliminary perspectives that had arisen.

### Data management and analysis

Verified transcripts were imported into NVivo 12 software (QSR International Pty Ltd, 2014) to manage and organize the data. Data analysis was conducted iteratively throughout the study using a thematic approach (Braun & Clarke, 2006; Fereday & Muir-Cochrane, 2006). A team based approach of thematic analysis was employed (Ekusai-Sebatta et al., 2021). Three of the authors (DES, DKM, and ESM) developed a codebook using both themes set a priori based on the literature review and survey outcomes from the quantitative study. Initially two team members (DES, ESM) coded a sample of the data independently to come up with more codes. Once these were reviewed together with the senior social scientist (DKM) a final codebook was developed which guided the rest of the coding process. The team read all transcripts to familiarize, mark and memo the data. Where differences emerged among the independent coders, they were solved by consensus. A thematic content analytical approach (Braun & Clarke, 2006) was employed to generate emergent themes and interpret the results and make comparisons. Findings were supported by representative quotes.

### Ethics approval

Ethical approval was obtained from the Makerere University School of Biomedical Sciences Higher Degrees and Research Ethics Committee (SBSHD-REC 517) followed by clearance by Uganda National Council for Science and Technology (SS 4490). Written informed consent was obtained from all participants prior to interview. All recordings and transcripts were de-identified, assigned special codes and stored on a password-protected computer. No participant identifying information was published.

## Results

There were 15 interviewees, the majority of which were male (12/15), and all were involved in international collaborative genomics and/or biobanking research. Five of the interviewees were clinical researchers, six were clinical epidemiologists and three were basic genetic scientists. Only one interviewee had formal training in clinical genetics. Participants had on average participated in research for a period of 12 years (SD 1.2, range: 3-22 years).

There were four themes: 1) opinions on the collection of biological samples for reuse; 2) opinion on biological sample export and regulation of collaborative biobanking research; 3) challenges faced by local researchers in biobanking research; and 4) possible solutions to improve/realize outcomes of biological sample/data sharing. The dataset to this work is accessible (Mafigiri, Ekusai, Munabi, & Mwaka, 2022).

### 1. Opinions on biological sample collection and storage

#### Leverage limited resources to advance knowledge and build laboratory capacity

Genetic and genomic researchers in Uganda expressed positive attitudes towards sample storage noting that the process was not only important for scientific advancement but could also contribute to capacity building as one of the elements to improve laboratory services in Uganda. Notably, biological sample storage was considered as an opportunity to leverage limited resources, which do not enable sustainably collecting biological samples in real-time. Researchers considered biological sample storage as an opportunity to provide a cohort of samples for future research that would otherwise be difficult to obtain if they were to rely solely on ‘real-time’ sample collection. Those who noted resource constraints in producing real-time sample collection felt that biological sample storage offered a cost effective, time saving yet informative option to efficiently conduct tests while at the same time build the capacity of local laboratory services. Overall, researchers acknowledged that there is limited local capacity for genetics research in Uganda.

> *We have to accept that our capacity is limited and even where we have capacity the costs can really be high. So, it might be more cost effective to just ship and analyze them* [the samples] *and we can’t deny that many times this is collaborative effort so the roles can be dispersed but it’s up to the institute, the sponsor, the principal investigator*. (R15, Female)

#### Adopt biobanks for efficient management

Researchers noted that establishment of biobanks was necessary to efficiently manage biological sample storage as a knowledge advancement and capacity building endeavor. Many discussed the feasibility of biological sample storage in LMIC in terms of documentation, storage capacity and governance as major categories that needed to be focused on to fully realize the potential of biobanks. Important issues raised to establish and improve biobanks included establishing documentation processes that were clear, secure, trustworthy and well governed. It was noted that there should be specific locations designated to host biobanks including repositories and centralized laboratory systems and venues. In terms of venues, participants mostly recommended that academic/research institutions were best placed to house the biobanks compared to public or private hospitals, particularly aimed at guarantying good governance.

#### Observe and be sensitive to cultural context of biological sample storage

Importantly researchers felt that biological sample storage also needed to be sensitive to the cultural context of the communities where the samples were being obtained. Respecting cultural values attached to biological samples was regarded as an important element that needed to be considered during collection and storage as well as when setting up biobanks. Some researchers were aware that not all biological sample types to be collecred were necessarily socially acceptable to the communities, let alone be stored for future unknown purposes. For instance, some researchers noted that collection of body parts like hair or finger and toe nails within some regions of Uganda was likely perceived as a taboo and not feasible as a scientific research procedure in the area as noted in the narrative below:

> *“Okay, culturally we believe that when somebody takes your hair and nails… then they have your life in their hands, they can decide to do anything with it …. So as believers in culture, if somebody collected my samples here and as they were taking them to a lab and may be somebody gave the lab runner something and they took a bit of hair and the nails and that is me …. they can control whatever they want …. and those are the beliefs. …we have actually had participants express so [no to hair sample] so we dropped it…. I am really not comfortable, yeah hair and nails no. So that’s why we dropped that particular study”. (*R03, Female)

#### Ownership of stored samples and assocaited data

Another important element discussed was the ownership of stored biological samples and associated data. Researchers felt that it needed to be clarified in the national ethics guidelines on who owned the stored samples. Notably, at the time of data collection there were guidelines from the Uganda National Council for Science and Technology for sample storage and data sharing (UNCST, 2014). However, participants’ limited knowledge about them could be a testimony of how imprecise they are on these matters. Researchers felt that there were lots of implications regarding ownership of biological samples such as when a discovery was made or when continuous use was needed, and how permission and consent processes would unfold. Most researchers seemed to be unsure of the rightful owner of the stored biological samples. Some researchers discussed about collaborative ownership that included combinations where the study sponsors, participants, researchers and regulatory bodies all had certain levels of influence and therefore single ownership would be difficult to determine. Others indicated that the samples belong to the study sponsor/funder because of their financial investment in the research. However, some researchers rightly stated that the sample donor retains ownership of the donated sample; with the Ugandan institution collecting the samples being a custodian as illustrated below:

> *… the bio-bank is a custodian. Simply a custodian. They don’t own these samples but in terms of ownership of the sample the sample belongs to the patient. But the patient offers this sample to the researcher…. Over the period of time and depending on the type of consent the patient still has control over that sample*. (R10, Male)

#### Ensure comprehensive consent processes exist

Related to being sensitive to the cultural context of biological sample storage, researchers also discussed the need to ensure that comprehensive informed consent processes were undertaken for biological sample storage. Researchers were keen to note that not many study participants or community members really knew what the process of biological sample storage involved. For instance, issues of ownership of samples, access to outcomes or rewards from the advancement of knowledge were hardly discussed in current informed consent processes for biological samples that were being stored. Researchers emphasized the need for honesty and transparency during the informed consent process.

#### Transfer/export of biological samples across borders advances knowledge by increasing access to technology

Another important issue that researchers discussed concerned was the transfer of biological samples out of the country of origin to the western world. Researchers noted that it was so common for the biological samples to be exported in part due to limited in-country technological capacity for sample storage and analysis. As such participants noted that it was good for the advancement of knowledge given the limited access to the technology needed to manage the samples or tests in most LMICs. However, biological sample storage in the Global North was reported to limit local capacity building efforts like infrastructure and human resource development. But the limited capacity was considered a longstanding problem that needed to be addressed to reduce the need for export of biological samples. Many researchers decried the decades-long practice of transferring samples out of Uganda, with some noting in no uncertain terms how it was an unfair practice to the development of the country.

> *“It is bad because it limits capacity building here, back at home”* (R05, Male).

#### Negotiate meaningful material transfer agreements (MTA)

Researchers appreciated the importance of well negotiated MTAs in the transfer of biological samples across borders. They however noted that many local institutions lack the bargaining power to negotiate collaborative agreements that are favourable to their interests. They also expressed concern about the lack of mechanisms for monitoring the execution of the MTAs. Researchers did not trust that the provisions of the MTA would be respected and upheld by collaborating scientists once the samples are shipped. Researchers posited that poorly negotiated MTAs are detrimental to local capacity building initiatives as was stated by this researcher:

> *So, the institutions are left on their own even when the guidelines are saying these samples should not be taken because we have local capacity. They are just signing the MTAs; the MTA is killing local capacity. So that is a very important document for us which needs to be taken well care of. (R09, Male)*

### 2. Opinions on sharing of biological samples and research data

#### Biological sample and research data sharing advances knowledge and collaborative research

Researchers perceived sharing of biological samples and data to have positive implications for research. They reported that biological sample sharing would enable advancement of knowledge and collaborative research as researchers who are unable to conduct primary data collection would have access to secondary data to apply their analytical skills as illustrated below:

> *… open access model ends up in public databases and the DBAC* [Data and Biospecimen Access Committee] *goes ahead to define how long it should take for the data to be available in public databases in the sense that how long will the investigator take to exhaust what he wanted to do with this data before throwing it out there in the public database. I think that the H3Africa model presents with a very good model in terms of international sharing of samples and data, and I think that’s the point where we should look*. (R12, Male)

Notably this observation was made in the period prior to the COVID-19 pandemic which more visibly revealed the possibility of inability to conduct primary data collection as an internationally-based researcher. However some researchers perceived that biological sample and associated data sharing often resulted into disproportionate rewards for local scientists in LMICs where the primary data was collected. They indicated that the varying (often limited) data analysis and writing skills meant that local scientists who contributed the most in generating primary data often gained the least from the outcomes of the research including particularly in the area of graduate training, publications and other awards.

> *Some of these collaborators ship because a sample is an asset, it brings in PhD students and funding. It can be used to leverage many things. I can use it to leverage NIH* [National Institutes of Health] *funding. I can use it to get more PhD students in my lab to collaborate with a certain lab, I tell you, if you want this, I give you my samples, you give me some students that will work on this or a pharmaceutical company, it’s an asset. It’s a value, now I think we need to start seeing it that way*. (R01, Male)

#### Local-global, cross-cultural authorship disputes

Biological and research data sharing was also reportedly rife with authorship disputes partciularly between LMICs and western counterparts. Some researchers noted that they either knew of or had personally experienced authorship disputes ranging from outright omission to position in the authorship list, with most valuable outcomes favoring their collaborators from the Global North as illustrated below:

*…massive tissue was exported and in the beginning, they were being exported for one thing, but subsequently they been analyzed and research papers have been published. So, what happens, is you just go to a conference and realize that the tissue came from Uganda; and the first form of annoyance is that there is no Ugandan attached to it as if they did not contribute to this at all. Secondly, if they are published without any regard to what is happening in Uganda; and the third is there is often no direct benefit to the society in Uganda, to the institution or even the people who collected this information and right now there is no protection*. (Female researcher)

### 3. Challenges faced in biological sample sharing

#### *Lack of effective means of tracking and monitoring* exported biological samples

Researchers noted the lack of a uniform regulatory system that applies to human biobanks used for genetic research purposes. They perceived this limitation to cause considerable variation in both national and international law that applies to use of biological samples, particulalrly DNA, personal health information and medical records across LMICs and their international collaborators. They expressed dismay about potential situation where researchers collaborating internationally may be operating unlawfully if they share research data and biological samples across borders where different laws are in operation. Similarly, many researchers expressed a fear of losing control of samples that are exported and stored away from their physical location of operation such as in overseas laboratories.

#### Inequity in sharing benefits for LMIC researchers dumpen morale and skills building

Researchers pointed out that there should be fair and equitable sharing of the benefits of research with individual participants and research communities. They however pointed out that oftentimes this is not possible because the available ethico-legal frameworks are unclear on this issue. Researchers further noted that lack of standardised guidelines for biological sample and data sharing inhibit cooperation among researchers. They attributed the lack of guidelines to limited capacity on the part of LMIC institutions and human resources as well as the over reliance on international collaborating partners to plan and implement most technical aspects such as laboratory based researches. They noted that the current status of biological sample and data sharing leads to inequity and unfair sharing of benefits for LMIC researchers and dumpens efforts to build capacity.

> *I think what we haven’t covered is about the issue of benefit sharing. We mentioned it but we didn’t discuss it fully. I personally think that communities should directly benefit from the benefits of genomic research. Study communities should be part of the patents in genomic research, and I think negotiations should involve them right from the beginning up to the end. We know that genomic research is beneficial, we know that there will be a lot of intellectual property and we believe that because the genes belong to these people, they should be respected in the sharing of these* [benefits]. (R14, Female)

#### Limited understanding of the regulations governing biological sample and research data sharing

Researchers were unanimous regarding the necessity for good biobank governance, especially in international collaborative research. However, they pointed out that many local researchers are not conversant with the guidelines and regulations that govern biological sample and data sharing in Uganda. They also noted that at times differences in regulations across international borders was an impediment to biological sample and research data sharing. They emphasized the need for harmonization of biobanking regulations across international borders with cognizance of socio-cultural considerations.

> *Nobody has permission to export data out of the country without getting permission from the solicitor general* [Government of Uganda lead representative on legal matters]. *I am reading from the current laws of the country, so researchers are currently carrying out an illegal activity of exporting data out of the country without the due release from the judicial authorities. Ahh so currently MTAs are one sided, they are legalistic favouring the collaborators. I think to me we should improve the collaborative frameworks around to show that our institutions here and communities and nations are more respected*. (R15, Male)

### 4. Possible solutions to improve/realize outcomes of *biological* sample/data sharing

#### Strengthen consent processes to improve understanding of biological sample storage

Researchers noted that strengthening the informed consent process would enhance potential participants’ understanding of biological sample storage and sharing processes. They noted that consent processes need to be more elaborate and should allow more time to engage in innovative ways to ensure that research participants learn about complex issues especially those related with genetics and genomics research. It was observed that conventional approaches of seeking informed consent may not be adequate when the subject of study is relatively novel to the local cultural context as is the case with biological sample storage in Uganda.

#### Empower LMIC researchers to negotiate MTAs and data sharing agreements

Participants noted that LMIC researchers and institutions needed to be empowered to negotiate MTAs and data sharing agreements that would enhance the possibilities to gain sufficient reward from the research process. Whereas researchers were cognizant of the limitation that funding comes from the Global North and thus the power to negotiate, they still felt that sometimes collaborators from the Global South were either not aware of the potential to negotiate for more meaningful and rewarding agreements.

#### Strengthen regulation of biological sample storage and transfer

Researchers also highlighted the need to strengthen regulation of biological sample storage and transfer processes in LMICs. At the time of data collection for example, whereas there were national ethics guidelines for regulating sample collection, storage and sharing; they were (national ethics guidelines) perceived to be ambigious and not clear about many potential ethical and social implications. It should be noted that efforts to strength such a framework have since been initiated by the UNCST. However participants noted that while it was being discussed and developed they felt that the framers were severely compromised given the fact that they heavily depended on funding from the Global North and thus needed to take into account their views with more favorability than would otherwise have been the case. Several participants also noted that the current national ethics guidelines are silent on the role of Research Ethics Committees in negotiating material and data transfer agreements. Researchers therefore recommended that UNCST should play a more active role in tracking and monitoring of exported samples by developing regulatory policies and empowering RECs to ensure that the terms and conditions of the MTA are respected by all parties. They opined that RECs should be given a clear mandate to review and approve MTAs and data sharing agreements.

#### Infrastructural and human resource capacity strengthening

Researchers pointed out that there is a need for capacity strengthening within LMICs to reduce the necessity for transfering biological samples while also building capacity for conducting sophisticated research procedures. They indicated that there should be deliberate effort to encourage foreign collaborators and funders to invest in local infrastructure and human resource capacity strengthening instead of exporting biological samples. They suggested that local scientists should also be encouraged to utilize the exported biological samples to acquire new knowledge, techniques and skills they can transfer and utilize when they travel back home.

> *Capacity is lacking here, they should be encouraged to come and develop capacity here such that instead of us sending the samples to them. They can come and do the research here and, in the process, build further capacity here to do this research either in terms of getting facilities or in terms of training people to do that sort of research*. (R08, Male)

## Discussion

The exponential growth of genetic and genomic research, including in LMICs like Uganda provides tremendous opprotunities for scientic advancement for the health and wellbeing of communities (Matovu et al., 2014; Parker & Kwiatkowski, 2016). This growth has however not been without some limitations including the unequal benefits for the Global South compared to the Global North. This inequity has been in part due to the technological gaps in laboratory capacity, sample storage, biobanking and future use of samples. However the recognition to reduce this inequity has grown as manifested by the H3Africa initiatives. Our study sought to examine the perceptions of genetic and genomic researchers towards the current practices of biological sample collection and storage for future use, and transfer particularly in the context of international collaborative research.

We noted that whereas there are lots of concerns about the current trends of sample collection, storage and transfer, researchers were cognizant of the need to not hamper knowledge generation given the limited capacity in LMICs. However, they were also keen to emphasise the need for capacity building within the LMICs. Just as other previous studies have highlighted the limitations of current practices of sample storage and processing from the Global North (Goisauf et al., 2019; Simeon-Dubach & Henderson, 2020), our study also highlights these concerns to still be at the heart of genetic and genomic researhers. Our study further points out the need to continue addressing the causes of such limited capacity to store, govern, and reuse biological samples on the continent.

Our study points out that whereas there are numerous guidelines that have been developed to guide international biobanking research, there remains cross-border variation in practices which impact biological sample sharing and transfer. Our participants emphasized the need for harmonization of biobanking regulations across international borders with cognizance of socio-cultural considerations. Biobanking research is rapidly evolving, therefore potential research participants and investigators need to be engaged more intensily to ensure that they are abreast with the current trends in biological sample storage and transfer. We also noted that there was limited capacity in negotiation of meaningful MTAs and biobanking processes which may continue to propagate the inequities currently being experienced between the Global North and South in terms of genomic and genetic research outcomes.

Empirical evidence underscores the importance of trust and governance in the success of biobank research. Our participants did not trust that the provisions of MTAs would be respected and upheld by collaborating scientists once the biological samples are shipped. This issue of distrust and antagonism towards biobanking is not uncommon (Broekstra, Aris-Meijer, Maeckelberghe, Stolk, & Otten, 2022; Matandika et al., 2020; Singh, Cadigan, & Moodley, 2022). Governance frameworks for biobanking research should aim to foster mutual trust between participants and researchers and institutions to whom samples are being donate (Wallace & Knoppers, 2012); between researchers in LMICs and other international collaborators (Munung et al., 2018); and between regulators and research implementors (Staunton & De Vries, 2020). The success of biobanking research is majorly dependent on people’s willingness to donate samples and their trust in responsible conduct of research and handling of their biological tissues.

Our findings suggest that there is a need to be sensitive to the cultural context of the community during the collection and storage of biological samples, as well as when setting up biobanks. Consent for biological sample storage and reuse should be culturally appropriate and should take cognizance of cultural sensitivities around the use of various types of biological materials as noted elsewhere (Tindana et al., 2014). There are several biological samples that are held sacred by some communities in Africa (Nguyen & Wong, 2006; van Bogaert & Ogunbanjo, 2008) and their collection and storage may be socially unacceptable, as such, care should be taken to ensure that the research intensions are not misconstrued by such communities. Further, the most commonly used informed consent approaches in biobanking are premised in Western-European world-view (Beauchamp & Childress, 2013). However, these approaches pose a challenge to informed consent for biobanking research in Africa because of the communitarian nature of society, customary beliefs, spirituality and relational autonomy (Akpa-Inyang & Chima, 2021). Therefore, ethical conduct of research in these settings requires culturally contextualized informed consent processes.

## Conclusion

Researchers acknowledged that there is limited local capacity for genetics and biobanking research in Uganda. However they expressed positive attitudes towards biobanking. They posited that biobanking offers a cost effective, time saving yet informative option to efficiently utilize biological samples while at the same time building local capacity. The importance of respecting cultural values in biobanking research was emphasized. Most researchers seemed to be unsure of the rightful owner of the stored biological samples yet the national ethics guidelines are clear on this issue. This was attributed to the limited understanding of the ethico-regulatory frameworks governing biological sample and data sharing in Uganda. Researchers appreciated the benefits of cross-border sharing of biological samples and research data but expressed displeasure with the practice of biological samples export. They felt that this practice was retrogressive and had adversely affected local capacity building efforts. Researchers felt that collaborative biobanking research between the Global North and south was not level because of poor biobank governance and stewarship, and the unfair and inequitable benefit sharing. There is a need for fair and equitable international collaboration where researchers from LMICs are treated with respect and as equal partners.

### Best practices

Participants appreciated the benefits of cross-border sharing of biological samples but decried the decades-long practice of transferring biological samples out of Uganda. They felt that this practice has adversely affected local capacity building efforts. They appreciated the importance of MTAs in collaborative biobanking research, however they noted that many local institutions lack the bargaining power to negotiate collaborative agreements that are favourable to their interests. In this regard, they raised concerns on the unfairness and inequity in benefit sharing in biobanking research in LMICs. Researchers also cited the lack of effective means of tracking and monitoring exported biological samples. These challenges are not unique to Uganda, a recent study in South Africa also reported discrepancy in governance processes for biobanking research including challenges with sample and data sharing, and inadequate approaches to benefit sharing and return of results (Singh & Moodley, 2021). They have also been associated with poor community engagement (Tindana et al., 2015; Tindana, Molyneux, Bull, & Parker, 2017) and inconsistency in defining ownership and custodianship of biological materials (Singh et al., 2022). Our findings suggest that fostering ethical biobanking research requires enhancing of informed consent processes, empowering researchers to negotiate MTAs and other associated collaborative agreements, strengthening biobank governance and local capacity strengthening.

### Research agenda

Our findings have revealed several concerns that may be impacting the sharing of biological samples and associated data in international collaborative research. However, these findings may not be generalizable to the larger research community in Uganda. There is a need for larger quantitative studies involving various stakeholders on the facilitators and barriers of effective biobanking collaborative research in low resource settings. Such information will provide an evidence base for better biobank governance and practice in these settings.

### Educational Implications

Our findings suggest that there is limited understanding of the ethico-regulatory requirements for the sharing of biological samples and associated data in collaborative biobanking research. For example, several researchers seemed to be unsure of the rightful owner of the stored biological samples yet the national ethics guidelines are clear that sample donors retain ownership and the collecting institution is just a custodian in trust. Other studies too have reported inconsistencies in understanding of ethical considerations in biobanking (Pawlikowski, Sak, & Marczewski, 2010; Singh & Moodley, 2021). There is a need for sensitization and on-going educational efforts to improve understanding of biobanking among researchers, research regulators and the community.

## Data Availability

All data produced in the present study are available online at https://doi.org/10.6084/m9.figshare.20367717.v1

## Declaration of conflict of interest

The author declare no potential conflicts of interest with respect to the research, authorship, and/or publication of this article.

## Sources of support

Research reported in this publication was supported by the National Human Genome Research Institute (NHGRI) of the National Institutes of Health under Award Number U01HG009810.

## Acknowledgement

We are thankful to Mr. Godfrey Bagenda, Ms. Deborah Ainembabazi, Prof. Joseph Ochieng and Dr. Janet Nakigudde for their various contributions to this work. We would also like to thank Prof. Nelson Sewankambo for his unwavering support and mentorship. Lastly, we would like to sincerely appreciate our study participants. The content is solely the responsibility of the authors and does not necessarily represent the official views of the US National Institutes of Health.

## Notes

### Competing Interest Statement

The authors have declared no competing interest.

### Funding Statement

This study was funded by the National Human Genome Research Institute (NHGRI) of the National Institutes of Health under Award Number U01HG009810.

### Author Declarations

Ethics committee of Makerere University School of Biomedical Sciences Research Ethics Committee gave approval for this work

